# Psychological resilience mediates the relationship between perceived neuropsychological impairment and quality of life in a sample of patients with multiple sclerosis

**DOI:** 10.1101/2022.08.13.22278745

**Authors:** Yunier Broche-Pérez, Rodneys M. Jiménez-Morales, Laura Ortiz Monasterio-Ramos, Johana Bauer

## Abstract

The impact of subjective cognitive concerns (SCCs) on the quality of life (QoL) of patients with multiple sclerosis (PwMS) has practically not been studied. In this study the relationship between subjective cognitive concerns and quality of life in PwMS was explored. Furthermore, to explore whether psychological resilience acts as a mediator in the relationship between SCCs and QoL. A total of 214 PwMS were surveyed using the Multiple Sclerosis Quality of Life Inventory, the Multiple Sclerosis Neuropsychological Questionnaire (MSNQ) and the Connor-Davidson Resilience Scale. Our results showed that, SCCs is a predictor of levels of perceived QoL in PwMS. Patients who report higher scores on the MSNQ also showed a worse quality of life in global terms. The results also showed that resilience mediates the relationship between SCCs and QoL, both for the physical dimension of quality of life (physical health composite) and for the mental health dimension (mental health composite). In our patients, as resilience levels increase, the negative impact of SCCs on QoL decreases. Considering that resilience is a modifiable protective factor, the implementation of interventions aimed at enhancing resilience can have a favorable impact on the psychological well-being and quality of life of patients with multiple sclerosis.

## Introduction

Cognitive problems are common in patients with multiple sclerosis (PwMS). Approximately 50% of PwMS present some type of cognitive impairment (CI), with learning problems, information processing speed, visual spatial skills, memory, and executive functioning being frequent (Chiaravalloti & DeLuca, 2008; Grzegorski & Losy, 2017; Jongen et al., 2012; Lyon-Caen et al., 1986; Rao et al., 1991). Several studies have shown that these cognitive alterations have a negative impact on the quality of life (QoL) of patients with multiple sclerosis (Benedict et al., 2005; Benito-León et al., 2002; Cutajar et al., 2000), undermining the realization of activities of daily living.

However, the impact of subjective cognitive concerns (SCCs) on the quality of life of PwMS has practically not been studied (D’Hooghe et al., 2019; Kletenik et al., 2019; Samartzis et al., 2014). According to Mitchell (2008), SCCs are a self-reported perception of dysfunction in cognition with or without impairment on objective cognitive testing.

Most of the research carried out on SCCs in PwMS has focused on exploring the relationship between subjective cognitive concerns and objective cognitive alterations. In this sense, while a group of investigations have reported the absence of a relationship between SCCs and objective cognitive alterations (Golan et al., 2018; Jougleux-Vie et al., 2014; O’Brien et al., 2007), other studies have suggested that patients with more SCCs also show worse objective cognitive performance when assessed with standardized cognitive tests (Henneghan et al., 2017; Marrie et al., 2005; Riccardi et al., 2021). Additionally, several studies have verified that the PwMS that report more subjective cognitive concerns also present, in general terms, higher levels of depression and anxiety (Benedict et al., 2003; O’Brien et al., 2007; Sonder et al., 2012).

The few studies that have explored the relationship between the impact of subjective cognitive concerns (SCCs) and the quality of life of PwMS show that patients with high levels of SCCs also present a greater deterioration in overall quality of life. For example, Samartzis et al. (2014) conducted a study on a sample of 100 PwMS with the aim of examining the e□ects of perceived cognitive dysfunction (using the Perceived Deficits Questionnaire (PDQ)) and of depression, on self-reported QoL. According to the results obtained, perceived planning/organization and perceived retrospective memory dysfunction predicted QoL scores in the patients, independently of the severity of disease and the severity of depression (Samartzis et al., 2014).

Another study conducted with a large sample of PwMS (n=751) had among its objectives to describe the relationship between SCCs and health-related quality of life (HR-QoL) (D’Hooghe et al., 2019). To assess SCCs, the authors used the MS Neuropsychological Screening Questionnaire (MSNQ), concluding that an increase in perceived neuropsychological impairment significantly contributed to explain low levels of HR-QoL. This relationship remained significant even after correction for sex, age, education and depression (D’Hooghe et al., 2019).

On the other hand, it is essential to explore which variables act as mediators of the relationship between SCCs and quality of life in PwMS, since, if modifiable, they could become potential therapeutic targets. In our opinion, a variable of great importance that we must consider in this scenario is psychological resilience. Psychological resilience is a process in which individuals display positive adjustment, despite being faced with significant adversity (Gallo & Bisecco, 2021). Recently, the controversy over the role of resilience in PwMS has increased (Gallo & Bisecco, 2021; Leavitt & Kever, 2021; Ploughman, 2021), with recent studies reporting that PwMS with higher levels of resilience may present better global disease-related outcomes, compared with patients showing low levels of resilience (Klineova et al., 2020). If, as we anticipated, resilience acts as a significant mediator between SCCs and QoL, the result could have psychotherapeutic implications to enhance quality of life in PwMS (considering that resilience is a modifiable protective factor (Alarcón et al., 2020)).

Therefore, this study has two main objectives. First, to explore the relationship between SCCs and quality of life in PwMS. Furthermore, to explore whether psychological resilience acts as a mediator in the relationship between SCCs and QoL in a sample of PwMS.

## Methods

### Study Design and Participants

An online cross-sectional study was carried out between August 13 and November 13, 2021. Patients with MS from 11 countries (Argentina, Mexico, Dominican Republic, Chile, Spain, Cuba, Colombia, Uruguay, Paraguay, Peru, and El Salvador) were invited to participate in the study through social networks (Instagram, WhatsApp and Facebook groups) and direct telephone calls were also made by their primary neurologists (using the personal information registered in the medical records). Several associations of MS patients also collaborated with the dissemination of the study. In total, 214 patients with multiple sclerosis completed the survey. This research is part of the project entitled

«Positive modulatory variables of perceived quality of life in patients with multiple sclerosis (ME-POSITIVE project)».

### Measures

Demographic and Clinical Information: The demographic variables explored included the age, disease duration, gender, education, marital status, country and MS phenotype.

Multiple Sclerosis Quality of Life 29 items version (MSQOL-29) (Rosato et al., 2016): The MSQOL-29 is a reduced version of the original 54-item inventory (MSQOL-54). MSQOL-29 consists of seven multi-item subscales: ‘physical function’; ‘sexual function’; ‘bodily pain’, ‘emotional well-being’, ‘energy’, ‘cognitive function’, ‘health distress’, ‘social function’, ‘health perceptions’, ‘overall quality of life’, and ‘change in health’. The scale also has two composite scores: physical health composite (PHC), and mental health composite (MHC).

The Connor-Davidson Resilience Scale 10 items version (CD-RISC-10) (Campbell-Sills & Stein, 2007): The 10-item CD-RISC was extracted from the original 25-item CD-RISC (Connor & Davidson, 2003). Patients’ rate items on a 5-point Likert scale, ranging from 0 (not true at all) to 4 (true nearly all the time). Total scores for the CD-RISC-10 rage from a minimum of 0 to a maximum of 40. Total scores are calculated by summing all 10 items. A higher score indicates higher resilience.

Patient-version of the Multiple Sclerosis Neuropsychological Questionnaire (p-MSNQ) (Benedict et al., 2003): The p-MSNQ is a 15-item self-report measure of perceived neuropsychological impairment (processing speed, dual task processing, attention, memory, executive function, and psychosocial comportment). The questionnaire items have 5 response options, 0 (does not occur) to 4 (very often, very disruptive). A total score is computed with a range from 0 to 60, a higher score indicates more cognitive problems. In this study, the Spanish version of the MSNQ validated in Argentina was used (Vanotti et al., 2009).

### Ethical statement

The ethics committee of the Department of Psychology of the Universidad Central «Marta Abreu» de Las Villas approved the study protocol. All procedures performed in this study were in accordance with the ethical standards of the 1964 Helsinki Declaration. Informed consent was obtained from all participants included in the study.

### Data analysis

The data were processed using SPSS/Windows (version 21). First, descriptive analysis was performed and an independent samples t-test was performed to compare psychological resilience, perceived neuropsychological impairment, and quality of life between relapsing-remitting and progressive phenotypes. Then, a Pearson’s correlation analysis was performed to assess the correlation between psychological resilience, perceived neuropsychological impairment and quality of life. Finally, a simple mediation analysis was performed using PROCESS macro (Hayes, 2018). We used bootstrapping sampling (n =5000) distributions to calculate the direct and indirect effects and confidence intervals (95%) of the estimated effects. Significance was determined when the confidence interval does not include zero. Before performing the mediation analysis, lack of multicollinearity, multivariate normality, and linearity were checked. The data was also checked for outliers.

## Results

### Characteristics of the sample

Tables 1 and 2 summarize the demographic and clinical information of the total sample (n = 214). A total of 179 patients (83.6%) presented the Relapsing-Remitting MS phenotype, while 35 patients (16.3%) reported a progressive phenotype (PPs; secondary progressive or primary progressive MS phenotype). In the sample, 168 participants (78.5%) were females, and most of the participants had a university-level degree (57.5%). The countries most represented in the study were Argentina (28.0%), Mexico (22.4%), Uruguay (13.1%) and Spain (10.3%).

**Table 1.** Participant demographic data (n=214).

| Demographic variable | Total (n=214)<br>n(%) | RRMS (n=179)<br>n(%) | PPs (n=35)<br>n(%) |
| --- | --- | --- | --- |
| Age | 39.84±10.80* | 39.89±10.80* | 46.14±11.73* |
| 18-35 | 76(35.5) | 70(39.1) | 6(17.1) |
| 36-50 | 94(43.9) | 77(43.0) | 17(48.6) |
| 51+ | 44(20.6) | 32(17.9) | 12(34.3) |
| Gender |  |  |  |
| Female | 168(78.5) | 148(82.7) | 20(57.1) |
| Male | 46(21.5) | 31(17.3) | 15(42.9) |
| Education |  |  |  |
| Primary education | 5(2.3) | 3(1.7) | 2(5.7) |
| Middle School | 23(10.7) | 18(10.1) | 5(14.3) |
| High School | 63(29.4) | 53(29.6) | 10(28.6) |
| University Degree | 123(57.5) | 105(58.7) | 18(51.4) |
| Marital status |  |  |  |
| Single | 83(38.8) | 70(39.1) | 13(37.1) |
| Married/Consensual union | 90(42.1) | 74(41.3) | 16(45.7) |
| Divorced | 40(18.7) | 34(19.0) | 6(17.1) |
| Widowed | 1(.5) | 1(.6) | - |
| Country |  |  |  |
| Argentina | 60(28.0) | 53(29.6) | 7(20.0) |
| Mexico | 48(22.4) | 38(21.2) | 10(28.6) |
| Uruguay | 28(13.1) | 25(14.0) | 3(8.6) |
| Spain | 22(10.3) | 17(9.5) | 5(14.3) |
| Dominican Republic | 21(9.8) | 16(8.9) | 5(14.3) |
| Cuba | 16(7.5) | 13(7.3) | 3(8.6) |
| Colombia | 9(4.2) | 9(5.0) | - |
| Chile | 4(1.9) | 3(1.7) | 1(2.9) |
| Paraguay | 2(.9) | 2(1.1) | - |
| Peru | 1(.5) | 1(.6) | - |
| El Salvador | 1(.5) | 1(.6) | - |

Note. \*(Mean ± Standard Deviation), EMRR (Relapsing-Remitting multiple sclerosis), PPs (Progressive phenotypes)

**Table 2.** Participant clinical and psychological characteristics (n=214).

| Variables | Total (n=214)<br>Mean ±SD | RRMS (n=179)<br>Mean ±SD | PPs (n=35)<br>Mean ±SD | t | p |
| --- | --- | --- | --- | --- | --- |
| Disease duration <sub>years</sub> | 7.06±7.67 | 6.64±7.22 | 9.21±9.47 | 1.826 | .069 |
| Quality of life (MSQoL-29) | 56.60±14.25 | 57.97±14.50 | 49.60±10.57 | 3.245 | **.001** |
| Physical Health Composite | 62.63±13.69 | 64.21±13.54 | 54.54±12.33 | 3.917 | **.001** |
| Mental Health Composite | 51.76±15.53 | 51.98±16.29 | 50.63±10.96 | .469 | .640 |
| Resilience (CD-RISC-10) | 26.72±8.19 | 26.31±8.34 | 28.80±7.11 | -1.648 | .101 |
| Fatigue (MFIS-5) | 10.85±4.43 | 10.66±4.50 | 11.80±4.02 | -1.387 | .167 |
| Cognition (MSNQ-SV) | 24.16±12.44 | 25.09±12.53 | 19.40±10.94 | 2.504 | **.013** |

Note. SE(Standard Deviation), EMRR (Relapsing-remitting multiple sclerosis), PPs (Progressive phenotypes)

Patients with progressive forms of the disease (secondary progressive or primary progressive MS phenotype) compared to RRMS patients showed significantly lower values in the quality of life variable. The differences were found in the total scores of the quality of life scale and in the physical health composite. In the case of the mental health composite, no significant differences were found between the groups.

On the other hand, RRMS patients reported significantly higher scores on the p-MSNQ scale compared to PPs patients, indicating higher levels of perceived neuropsychological impairment (PNI). The variables fatigue and resilience did not show significant differences between the groups.

### Correlation between variables

There was a positive and significant correlation between quality of life and resilience, suggesting that patients with higher levels of resilience also report a better overall quality of life and vice versa. Physical and mental health composites also showed a positive and significant correlation with quality of life scale scores. In addition, total score of the MSQOL-29 and its both composites (physical health and mental health composites) also showed a significant and inverse correlation with the p-MSNQ scale. According to these results, patients who report lower perceived cognitive functioning (higher scores on the p-MSNQ scale) also have a lower quality of life (lower scores on the MSQOL-29) (table 3).

**Table 3.** Correlation between variables

|  | 1 | 2 | 3 | 4 | 6 |
| --- | --- | --- | --- | --- | --- |
| 1. Quality of Life | - |  |  |  |  |
| 2. PHC | ,809** | - |  |  |  |
| 3. MHC | ,819** | ,434** | - |  |  |
| 4. Resilience | ,420** | ,245** | ,499** | - |  |
| 5. Cognition | -,372** | -,210** | -,402** | -,384** | - |

Note. \*\* Correlation is significant at the 0.01 level (2-tailed), PHC (Physical Health Composite), MHC (Mental Health Composite)

### Mediation analysis

To explore the mediating effect of resilience between perceived neuropsychological impairment and quality of life, three simple mediation analyzes were conducted. The first of them (figure 1a) considers the total scores of the quality of life scale (MSQOL-29) as the dependent variable. Furthermore, we explored the mediating effect of resilience on the physical health composite (figure 1b), and the mental health composite (figure 1c) independently.

**Figure 1.**
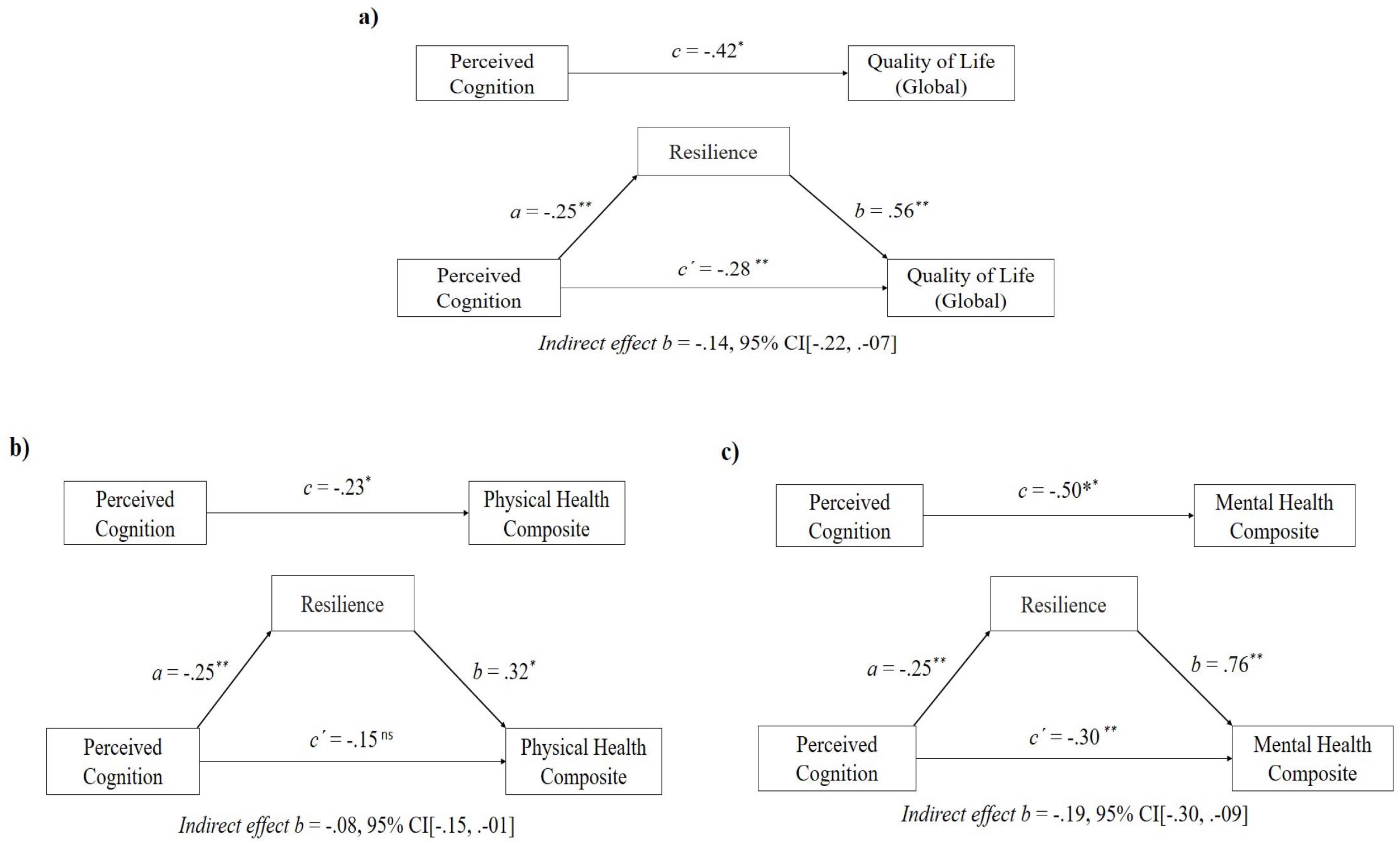
The result of simple mediational models, *p < .05, **p < .001, ns (no significance). Values shown are unstandardized coefficients.

As seen in figure 1a, the direct effect from PNI to psychological resilience (path a) was significant (β = -.25, 95% CI[-.33, -.17] t = -6.051, p < .001), showing that increasing levels of perceived neuropsychological impairment is related to a decrease in resilience levels in our PwMS. Additionally, the direct effect from resilience to quality of life (path b) was also significant (β = .56, 95% CI[.34, .79] t = 4.974, p < .001). When the levels of resilience increase, we can also predict an increase in the QoL in our patients. On the other hand, path c’ indicates that perceived neuropsychological impairment predicts QoL (β = -.28, 95% CI[.34, .79] t = -3.772, p < .001). According to this result when the levels of PNI increase, a decrease in quality of life levels can be predicted.

The total effect of perceived neuropsychological impairment on quality of life was significant (β = -.42, 95% CI[-.57, -.28] t = -5.835, p < .001), and when psychological resilience, which is the mediator, is included in the model, this effect (β = -.42) decrease (β = -.28), confirming the existence of a significant partial mediation (β =-. 14, 95% CI[-.22, .-07]). According to this result, psychological resilience acts as a mediator between perceived neuropsychological impairment and quality of life in our sample of PwMS.

Our results also showed (figure 1b) the direct effect from resilience (β = .32, 95% CI[.08, .56] t = 2.685, p = .007) to physical health dimension of quality of life (path b). In addition, it was found that the direct effect (path c’) of the perceived neuropsychological impairment on the physical health dimension of quality of life was not significant (β = -.15, 95% CI[-.30, .005] t = -1.897, p = .059). However, the total effect of perceived neuropsychological impairment on physical health dimension of quality of life was significant (β = -.42, 95% CI[-.57, -.28] t = -5.835, p < .001). In this sense, it was observed that when psychological resilience is included as a mediator in the relationship between perceived neuropsychological impairment and physical health dimension of quality of life, the total effect (β = -.23) decrease (β = -.15), verifying the existence of a significant mediation (β = -.08, 95% CI[-.15, .-01]).

On the other hand (figure 1c), resilience also showed a direct effect on the mental health dimension of quality of life (β = .76, 95% CI [-.46, -.15] t = 6.492, p < .001) (path b). It was also observed that the direct effect (path c’) of the perceived neuropsychological impairment on the mental health dimension of quality of life was significant (β = -.30, 95% CI[-.46, -.15] t = -3.953, p=.001). The total effect of perceived neuropsychological impairment on mental health dimension of quality of life was significant (β = -.50, 95% CI[-.65, -.34] t = -6.387, p < .001). The results indicated that when psychological resilience is included as a mediator in the relationship between the perceived neuropsychological impairment and mental health dimension of quality of life, the total effect (β = -.50) decrease (β = -.30), confirming the existence of a significant mediation (β = -.19, 95% CI[-.30, .-09]).

## Discussion

The purpose of this study was to explore the relationship between SCCs and quality of life in PwMS. In addition, whether psychological resilience acts as a mediator in the relationship between SCCs and QoL in a sample of PwMS was also explored.

First of all, our results showed that, in our sample, SCCs is a predictor of levels of perceived quality of life in PwMS. Patients who report higher scores on the MSNQ also showed a worse quality of life in global terms. The negative impact of SCCs on quality of life was significant both for the physical dimension of quality of life (physical health composite) and for the mental health dimension (mental health composite).

Our results support the conclusions of the study conducted by Samartzis et al. (2014) where they reported that the presence of SCCs (perceived planning/organization and perceived retrospective memory dysfunction) predicted QoL scores in PwMS, independently of the severity of disease and the severity of depression. In line with our results we can also mention a study conducted by D’Hooghe et al. (2019). In a sample of 751 PwMS, the researchers found that an increase in perceived neuropsychological impairment significantly contributed to explain low levels of health-related quality of life (HR-QoL), regardless of sex, age, education and depression.

It has also been suggested that SCCs, in addition to being related to a decrease in the quality of life of PwMS, could be are also related to brain volume changes. For example, in a study involving 158 PwMS, it was found that SCCs were associated with reduced cortical gray matter and thalamic volumes, regions of the brain that have been implicated in objective cognitive impairment (Kletenik et al., 2019).

However, there are also studies that have reported a weak association between SCCs and QoL in multiple sclerosis patients. In this case, an investigation carried out by Gold et al. (2003) where a weak association between SCCs and objective cognitive impairment and also between SCCs and QoL was reported. According to the authors, the existence of a low association could be related to the characteristics of the instrument used to assess SCCs in the study (HAQUAMS fatigue/thinking scale), which not only assess cognitive function but also includes items referring to fatigue. In this sense, a strength of our study was the use of a validated self-reported measure for perceived cognitive impairment (the p-MSNQ), which increases the validity of our conclusions (Benedict et al., 2003; D’Hooghe et al., 2019; Vanotti et al., 2009).

In addition, whether psychological resilience acts as a mediator in the relationship between SCCs and QoL in a sample of PwMS was also explored. The results showed that resilience mediates the relationship between SCCs and quality of life. In our patients, as resilience levels increase, the negative impact of SCCs on quality of life decreases. The mediating effect of resilience was significant both for the physical dimension of quality of life (physical health composite) and for the mental health dimension (mental health composite).

Although we are not aware of any previous study that explored a hypothesis similar to ours, there is evidence of the mediating capacity of resilience in PwMS. For example, it has been found that in patients who report high levels of resilience, there is a lower impact of secondary symptoms such as pain and fatigue on quality of life (Terrill et al., 2016). Recently it has also been reported that PwMS who showed high psychological resilience presented better objective performance on the MS Functional Composite (MSFC) and motor outcomes, especially gross motor function (i.e., grip strength, gait endurance) (Klineova et al., 2020).

There are several tentative explanations that could constitute mechanisms of action that explain the mediating effect that resilience has shown in our study. A possible mechanism could be related to a decrease in the levels of depression and anxiety in patients who show higher levels of resilience, which would explain a lower perception of SCCs and therefore a better quality of life. Regarding this hypothesis, there are previous studies that have confirmed that the version of the MSNQ self-report form (s-MSNQ) was significantly correlated with measures of depression and anxiety, but was not correlated with objective cognitive impairment outcomes, unlike the informant version of the MSNQ (i-MSNQ) that has shown demonstrating significant correlations with objective cognitive impairment, but not with measures of depression and anxiety (Benedict et al., 2003; O’Brien et al., 2007; Sonder et al., 2012). In this sense, there is evidence that resilience is a protective factor that reduces symptoms of anxiety/depression in PwMS, increasing quality of life regardless of the physical disability level (Nakazawa et al., 2018).

Another tentative mechanism of action that would explain our results is related to the cognitive reserve construct. The cognitive reserve hypothesis posits the existence of a group of factors that contribute to better cognitive coping against disease-related cognitive decline (Stern, 2002), which would imply that the impact of disease burden on cognitive status is stronger among persons with lesser reserve (Sumowski & Leavitt, 2013). In this sense, the patients in our sample with higher levels of resilience could exhibit a group of behaviors that act as enhancers of cognitive reserve, such as practicing more physical exercise, showing better coping with stress, performing a greater number of cognitive leisure activities (read books, produce art, play structured games, participate in hobbies, etc.), which would reduce the negative effect of MS disease burden on perceived cognitive status and increase quality of life. An example is a study carried out by Notario-Pacheco et al. (2014) where it was found that PwMS with high levels of resilience lived with less disability and fatigue, reported greater participation, exercised more, consumed a healthier diet and lived with greater social support and financial security, compared to the lower scoring group (Notario-Pacheco et al., 2014). In this way, psychological resilience could be considered as an “umbrella” proxy of the cognitive reserve. This mechanism is extremely valuable, fundamentally for the development of interventions aimed at increasing the quality of life in PwMS.

Our results must be analyzed considering some limitations. First, this is a cross-sectional study so it is difficult to accurately elucidate causal relationships between subjective cognitive complains, psychological resilience and quality of life. Secondly, the sample is made up mostly of patients who present the relapsing remitting phenotype (83.6%) and to a lesser extent by patients with progressive clinical forms (16.3%). It is well known that both RRMS and progressive MS present important differences in the severity of physiological, psychological, and motor symptoms; which impacts in different ways on the quality of life of these patients (Nag et al., 2021). In future studies it is also important to control for the effect of covariates such as depression, anxiety, EDSS and fatigue. These variables are key to understanding the relationship between psychological resilience, quality of life and cognitive functioning in PwMS (Arnett et al., 2018; Bol et al., 2010; Vissicchio et al., 2019).

## Data Availability

All data produced in the present study are available upon reasonable request to the authors.

## Acknowledgement

First of all we want to express our gratitude to all the people with MS who participated in the study. We especially thank the coordinators and patients of the EMA (Esclerosis Múltiple Argentina) and ALCEM (Asociación de Lucha Contra la Esclerosis Múltiple) associations, as well as the patient associations of Mexico, Uruguay, Cuba and the Dominican Republic. We also thank Rocio Seijas (Escleroamigos) and Fernando Champomier (Emstrongs) who from their projects supported our research from the beginning. Finally, we especially thank Nicolás Edgardo Costa, host of the radio program «El juego no termina» (ALCEM podcast series made by people living with MS).

## Data availability statement

The datasets generated for this study are available on request to the corresponding author.

